# Forecasting Blood Supply in Chinese Major Cities by Fractional Grey Prediction Model and Linear Regression Model

**DOI:** 10.1101/2023.04.25.23287469

**Authors:** Feng Lin, Xu He, Huifang Zhang, Zhong Liu, Yi Huang

**Affiliations:** Institute of blood transfusion, Chinese academy of medical science and Peking union medical college, No.26 Huacai Road, Chengdu, 610052, People’s Republic of China

**Keywords:** Blood Supply, Fractional Grey Prediction model, Linear regression model, Resident Population, GDP

## Abstract

**Background and Objectives:** The aim of this study is to predict the quantity of blood donations in major Chinese cities from 2023 to 2026. Blood is a scarce and perishable resource, and its availability in a particular area depends on the amount of blood donated in that area. Therefore, it is essential for healthcare providers and policymakers to have accurate predictions of future blood donation quantities in order to plan for transfusion treatments and operations. This study aims to provide insights into the future blood supply and allocation in major Chinese cities.

**Methods:** We used a combination of linear regression and fractional grey prediction models to predict blood donation quantities in major Chinese cities from 2023 to 2026. The models were developed based on historical data of blood donation quantity, resident population, and gross domestic production. We compared the predicted values from the models to cross-check the consistency of the predictions. When they were not consistent, the predictions based on the recent historical data were chosen. We cross-checked the predictions with historical data to assess their accuracy.

**Results:** We found that the predicted future donation quantity in major Chinese cities from 2023 to 2026 would be around 20 ± 3U per 1000 persons, and the models suggested that these predictions were credible in at least half of the cities examined. We also observed a slightly larger differential order of the grey prediction model in the cities of northeast China compared to the rest of the cities. Furthermore, we found that the reliability of the predictions varied, with over half of the cities showing consistent predictions and others showing inconsistent or unreliable results.

**Conclusion:** Our findings provide important insights for healthcare providers and policymakers to plan for blood supply and allocation in the future. However, the reliability of the predictions varied, indicating the need for updating historical data to improve the accuracy and reliability of prediction models. We also find that in northeast of China, the blood donation quantity might be bias from 20U per 1000 persons after 2026, according to the differential order of the fractional grey prediction model. Overall, our study highlights the importance of accurate predictions of blood donation quantities for effective healthcare planning and resource allocation.

## 1 Introduction

Blood transfusion services play an essential, underpinning role in health systems. All countries face challenges in making sufficient supplies of blood and blood products available and sustainable [1].Since the demand for blood in China exceeds the supply, it is necessary for the governments of the cities to forecast the blood donation quantity. China has been using voluntary and non-remunerated blood donation for more than two decades [2]. The country collects whole blood through 452 blood centers, which include 32 provincial, 321 regional and 99 county-level centers [3]. Moreover, since 2012, Chinese donation databases have maintained detailed and interlinked data on donors, donations, blood components and transfusions. However, due to the COVID-19 pandemic, the continuous records only go up to 2018. Now we need to predict the blood donation quantity for 2023-2026. If we have continuous time records of blood donation quantity up to 2021, we might have some methods to predict the blood donation quantity in the next few years. However, if we only have the records of blood donation quantity in the last few years and need to forecast the blood donation quantity in the future 3-5 years, we have to rely on other data.

Many approaches have been proposed to predict the blood donation quantity, such as neural networks [4,5] or time series analysis [6]. Our sample size of data is not large enough to use neural networks method. The strong volatility and stochasticity of monthly data of blood donation quantity makes it hard to predict the blood donation quantity by time series analysis. So we have to turn to the annual data. However, the small sample size of annual data makes it difficult to make an accurate prediction by time series analysis methods such as Autoregressive Integrated Moving Average, Exponential Smoothing, etc. Data from the World Health Organization (WHO) shows that blood donation is related to local economies; cities with larger populations and higher incomes tend to have higher blood donation rates [7]. This implies that the economy or population of a city or an area can be used to estimate the donation quantity. The statistic data of economy or population of a city or a province is also published once a year. To make accurate predictions based on the small sample size, we have to try grey forecasting models.

Grey forecasting models proposed by Deng (1982) [8], have been utilized successfully in prediction of time-series data with small sample size in many fields [9–11]. If our annual data follow the conditions of the grey sequence, it is possible to forecast the blood donation quantity by first order univariate grey model abbreviated as GM(1,1) [12, 13]. If the time series data of annual blood donation does not meet the grey sequence conditions, we may use the fractional grey prediction models. To cross-referencing the predicted blood donation quantity, we also use linear model to predict the blood donation quantity. Linear correlation analysis is a method of using the degree of correlation of analysis data to support or refute hypotheses. It can be used to detect whether there is an association between two or more variables [14, 15]. When the linear relation holds, the blood donation data can be represented by economy or population data with a linear regression model.

We will introduce the methods in details in section 2, and show the results in section 3. In section 4 we will comment on the our forecasting results by our method.

## 2 Materials and Methods

### 2.1 Ethical approval

All data involve in the study had given written informed consent by National Blood Safety Center. The current project was approved by the review board of the Research Ethics Committee of Institute of Blood Transfusion, Chinese Academy of Medical Sciences and Peking Union Medical College on April 25, 2023 (registration number 2023018).

### 2.2 Materials: data

Data on blood donation quantity in 2012-2018 were supplied by the Chinese donation databases. There are hundreds of cities in China. As the majority of Chinese people live in major cities, we can choose one or two of the major cities from each of provinces to represent them. The economic and donation volume are correlated with the local population size. In each of China’s provinces, blood donation in the provincial capital city is much greater than that in non-provincial capital cities. 36 major cities were chosen to represent each region of the Chinese mainland. Each province was represented by at least one city in its region. The levels of blood donation in these cities were the largest or the second largest in the provinces where they were located.

Data on gross domestic products (GDP) and the number of inhabitants in the 36 major cities each year in 2012-2021 were obtained from the Annual Statistical Review (http://www.stats.gov.cn/sj/ndsj/). Correspondingly, annual data on blood donation in these cities were chosen from Chinese donation databases. As there is only one major blood center in Hainan province, the whole of Hainan province is regarded as a single city (the area of Hainan is smaller than that of the city of Chongqing).

Readers must have notice that the time span of the blood donation data is inconsistent with those of resident population and GDP. Due to missing blood donation data for the year 2019, and the impact of the COVID-19 pandemic on the behavior of donors from 2020 to 2022, it has been difficult to accurately reflect normal blood donation levels. We select the historical data of blood donation from 2012-2018. The resident population and GDP are not affected much by the pandemic, so we had better take the actual data as recent as possible.

### 2.3 Methods:Regression model

As the data from WHO shows that the blood donation is related to local economies, we will examine whether there is a linear relationship between the economy and the blood donation quantity by calculating the correlation coefficients of blood donation quantity and the GDP. Additionally, the correlation coefficients of blood donation quantity and number of inhabitants in the cities will also be calculated. If the results show that the blood donation quantities and the GDP or resident population have a linear relationship, then it is possible to use a linear regression model to describe the blood donation quantities by the number of inhabitants or the GDP.

If the correlation coefficients between the blood donation quantity and other factors (such as GDP or number of inhabitants) are linear, the estimation of the blood donation quantity can be modeled by the linear regression model. Regression analysis is to construct mathematical models which describe or explain relationships that may exist between variables [16]. The distribution of blood donation quantity in the 36 cities is statistically the same as that of GDP or resident population, the variables of GDP or resident population can represent the blood donation quantity by a linear model as formula (1).

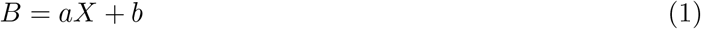

in which *B* denotes blood donation quantity, and *X* denotes the variable GDP or resident population depending on which variable we choose to represent blood donation quantity. The coefficients *a, b* are determined by the least squares method (LSM) [16]. When one variable can be linearly represented by another (for example, GDP and blood donation quantity in each of the cities), the 2 variables are in fact in the same linear space. This reduces the information of the undetermined variables.

The correlation coefficients of the blood donation quantity and the resident population are shown in table 1. The number 1.5447212172667432e-12 means 1.5447212172667432 *×* 10^−12^, and so on.

**Table 1:** The correlation coefficients of donation quantity and resident population

| Year | Correlation coefficients | P-values |
| --- | --- | --- |
| 2012 | 0.88005148005148 | 1.5447212172667432e-12 |
| 2013 | 0.892921492921493 | 2.485251209064001e-13 |
| 2014 | 0.885971685971686 | 6.849416607630513e-13 |
| 2015 | 0.8844272844272845 | 8.504240275126934e-13 |
| 2016 | 0.8913770913770913 | 3.1316283828092587e-13 |
| 2017 | 0.8885456885456886 | 4.742202059952288e-13 |
| 2018 | 0.9011583011583012 | 6.803778976419801e-14 |

The correlation coefficients of the blood donation quantity and the GDP are shown in Table 2. Table 1 and Table 2 show that the blood donation quantity has linear relationship with both the GDP and resident population. So the blood donation quantity in each of the cities can be represented by the variables such as GDP or resident population in each of those cities. The linear models of blood donation represented by the resident population from 2012-2018 are shown in Table3. Similarly, the linear regression models of the blood donation represented by the GDP in 2012-2018 are shown in table4. The errors of the linear regression models are the mean relative errors (MRE) which are defined as

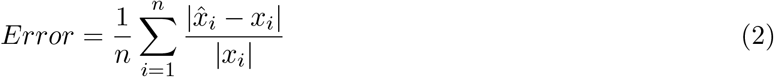

in which 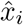 is the estimated value and *x*_*i*_ is the actual value. The relative errors for Huhhot and Lhasa are around or even exceed 100%. Therefore, we also report the MRE excluding these two cities in parentheses in Tables 3 and 4. As shown in the results, the mean relative errors are not very low. However we can take advantage of the large difference since the grey prediction model is also known to be inaccurate for long-term prediction. The large error difference might compensate for the error caused by the grey model. Therefore, we can combine both the grey prediction model and the linear model to obtain more reliable predicted values.

**Table 2:** The correlation coefficients of donation quantity and GDP

| Year | Correlation coefficients | P-values |
| --- | --- | --- |
| 2012 | 0.7817245817245818 | 1.8194655124655762e-08 |
| 2013 | 0.7786357786357788 | 2.251396340719785e-08 |
| 2014 | 0.8054054054054054 | 3.145353372430032e-09 |
| 2015 | 0.8005148005148005 | 4.606098874518366e-09 |
| 2016 | 0.8182754182754184 | 1.0929047445396942e-09 |
| 2017 | 0.8046332046332048 | 3.342980030097363e-09 |
| 2018 | 0.8010296010296011 | 4.427002183341357e-09 |

**Table 3:** The linear models of donation quantity (B) and resident population (R)

| Year | Linear model | $a$ in formula(1) | $b$ in formula(1) | MRE (%) |
| --- | --- | --- | --- | --- |
| 2012 | $B = 175.9 R + 41331.7$ | 175.90561077 | 41331.71676768 | 45.95 (22.72) |
| 2013 | $B = 175.9 R + 39763.6$ | 175.86355971 | 39763.55844214 | 44.66 (21.48) |
| 2014 | $B = 180.1 R + 40452.0$ | 180.07981082 | 40451.97951096 | 49.40 (22.99) |
| 2015 | $B = 188.4 R + 33427.2$ | 188.38971953 | 33427.2458664 | 43.19 (23.01) |
| 2016 | $B = 190.9 R + 38413.1$ | 190.91462657 | 38413.08498905 | 52.46 (22.27) |
| 2017 | $B = 192.4 R + 40488.1$ | 192.38416203 | 40488.05418258 | 50.00 (22.08) |
| 2018 | $B = 187.8 R + 47107.3$ | 187.82137441 | 47107.30507496 | 116.68 (23.73) |

**Table 4:** The linear models of donation quantity (B) and GDP (G)

| Year | Linear model | $a$ in formula(1) | $b$ in formula(1) | MRE (%) |
| --- | --- | --- | --- | --- |
| 2012 | $B = 21.0 G + 67846.1$ | 2.10178662e+01 | 6.78460695e+04 | 65.67 (30.68) |
| 2013 | $B = 19.3 G + 67399.1$ | 1.93438150e+01 | 6.73990749e+04 | 66.16 (27.81) |
| 2014 | $B = 19.1 G + 61651.8$ | 1.91994637e+01 | 6.16517846e+04 | 63.71 (28.02) |
| 2015 | $B = 18.6 G + 58293.8$ | 1.85994605e+01 | 5.82937604e+04 | 59.36 (29.24) |
| 2016 | $B = 16.9 G + 70935.9$ | 1.68723227e+01 | 7.09359219e+04 | 77.43 (29.72) |
| 2017 | $B = 15.2 G + 76728.7$ | 1.52178638e+01 | 7.67286558e+04 | 75.75 (30.67) |
| 2018 | $B = 13.7 G + 86721.2$ | 1.37154761e+01 | 8.67211713e+04 | 179.70 (32.98) |

So far we have introduced the approach for predicting the blood donation quantity in 2023-2026 by linear models. We will employ the grey prediction model to derive the future values of resident population and GDP in 2023-2026. However, this model has limitations for long-term predictions. It is more suitable for near-future predictions. If we had the blood donation quantity in 2019-2021, we could have built the linear regression models for blood donation in 2019-2021 which would have made the prediction more accurate. But the record of blood donation quantity is missing in the year of 2019, and in 2020-2022 the COVID-19 pandemic affected the blood donors’ behavior. This means that the blood donation quantity in 2020-2022 can not reflet the normal blood donation levels. So we use the linear regression models in Tables 3 and 4 to obtain the blood donation quantity in 2023-2026 as intervals.

### 2.4 Methods: Grey Prediction Model

Grey Model is a statistical method used to analyze and predict changes in a time series that have no clear trend or linear correlation [12, 13]. Grey sequence is a method of nonlinear analysis applied to time series data while grey model is a mathematical model used to predict future values. The methods were developed in the 1980s and have been used predominantly in engineering and economics. They have found applications in finance and other fields due to their robustness in handling time varying data. Grey sequence creates nonlinear relationships between consecutive values while grey model builds a forecasting model based on the cumulative sum of the Grey sequence. Both methods are commonly applied to short-term forecasting since they do not require a large amount of historical data and can quickly capture the underlying trend of a time series.

A single variable grey model is abbreviated as GM(1,1), and its’modelling procedures are briefly as follows [13] (Deng, 1989). Let 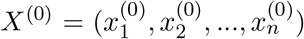 be an original sequence, that is, the elements of *X*^(0)^ are the observed values. All of the elements in *X*^(0)^ are required to be positive. The first order accumulative generation operation (1-AGO) sequences 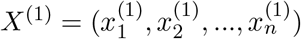 is defined as

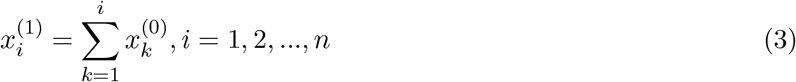

The mean neighboring generating sequence 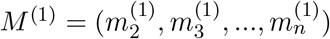 is defined as

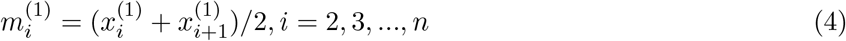

If all of the stepwise ratios *σ*_*i*_ of 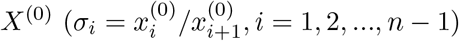 are in an interval, the sequence is called quasi smooth. A quasi smooth sequence can be embedded into a exponential sequence [17] which is a solution of an ordinary differential equation. Generally speaking, the interval of the stepwise ratios is set as (*e*^−2*/*(*n*+1)^, *e*^2*/*(*n*+1)^). When the sequence is quasi smooth, the GM(1,1) is defined as following

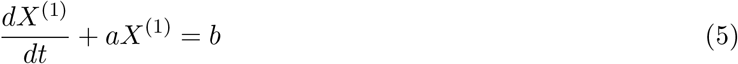

in which the parameters *a* and *b* can be determined by least squares method. The order of the elements in sequence *X*^(1)^ corresponds to the discrete time. So the model (5) is changed into a difference equation.

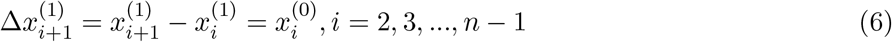

And 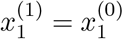. The parameters *a, b* form a vector

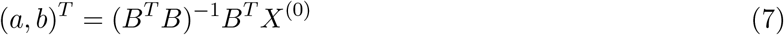

Where *T* means transpose and −1 means the inverse. The sequence *X*^(0)^ in formula (7) drops the first element 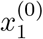. And the matrix *B* is

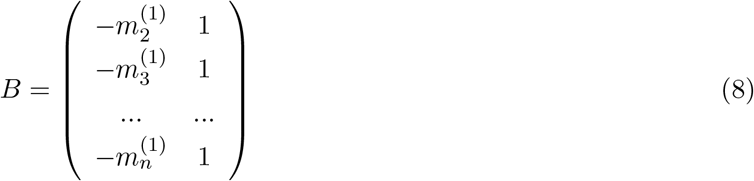

According to the general solution of ordinary differential equation, the elements of sequence *X*^(0)^ and *X*^(1)^ have the relations

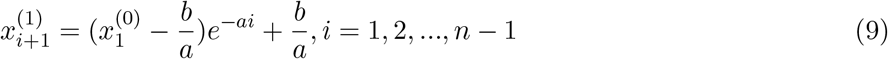

and

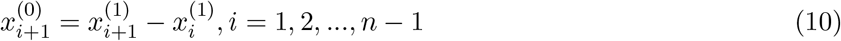

Using formulas (9) and (10) we can predict the future elements of sequence *X*^(0)^.

If the 1-AGO series *X*^(1)^ is smooth enough, GM(1,1) can predict the future elements accurately. However, the original sequence *X*^(0)^ may not always produce a smooth sequence *X*^(1)^. This means that the elements in sequence *X*^(0)^ have different weights. Generally speaking, the earlier elements have less influence on the later ones. The GM(1,1) model (5) is a first-order ordinary differential equation. The initial value determines all of the results. This means that the earlier elements have the same influence on the later ones, while the fractional differential derivative takes the memory effect into consideration.

In 2013, Wu et al [18] firstly introduced the fractional-order accumulation into the GM(1,1) model to further improve the accuracy of model. The fractional order *r* = *p/q* where *p, q* are integers which guarantees *r* to be a rational number. Increasing *r* can increase the weights of old data, which can put more emphasis on the older data. The elements in the accumulative sequence *X*^(*r*)^ are defined as

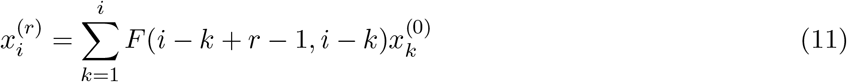

In which

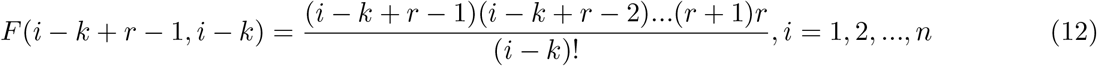

and *F* (*r* − 1, 0) = 1. When *r* = 1 the sequence *X*^(*r*)^ becomes 1-AGO. By definition [18], when *r* = 0, *F* (*i* − *k* + *r* − 1, *i* − *k*) = *F* (*i* − *k* − 1, *i* − *k*) = 0. *X*^(*r*)^ satisfies the first-order differential equation in the form as (5). Formula (12) can also be written in the form of Γ function

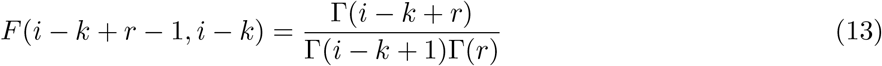

The elements 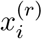 can be transformed into the 1-AGO sequence elements 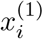 by

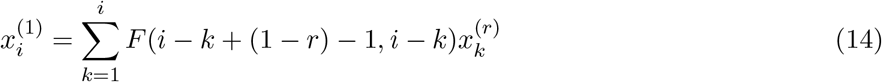

And 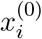 can be figured out by formula (10). The error of the estimated 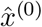 and the actual *x*^(0)^ is defined

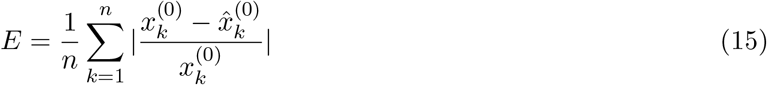

By the principle of new information priority [18], *r* ∈ (0, 1). The exact value of *r* corresponds to the minimal error in (15). We can calculate the formula (15) under several values of *r* from 0.05 to 0.9 and select the best *r* = *r*^*^ which corresponds to the minimal error.

### 2.5 Methods: Determination of Error Boundary for Evaluation and Determination

To assess the reliability of preliminary predictions from fractional prediction models or linear regression models, we must compare them with actual historical data. The latest historical records of blood donation quantity are from 2018. Because there is likely a bias between the 2018 data and data after 2020, we use the 2018 data as the reference point. This means that we regard the 2018 data as the midpoint of the reference assessment interval. To determine the upper and lower bounds of the reference assessment interval, we use historical data (2012-2018) on blood donation quantities. We calculate the average ratio of the range to the maximum and minimum donations across cities and present them in Table 5. In the column of range-to-minimum ratio, two cities have ratios above 1. We exclude these two cities and recalculate the average range-to-minimum ratio. The result is shown in parentheses.

**Table 5:** Average ratio of range to maximum or minimum donations across cities

| City | Ratio of range to minimum value | Ratio of range to maximum value |
| --- | --- | --- |
| Beijing | 0.212177857 | 0.175038552 |
| Tianjin | 0.154652645 | 0.133938675 |
| Shijiazhuang | 0.192380155 | 0.161341292 |
| Taiyuan | 0.307420289 | 0.235135015 |
| Huohhot | 0.268496946 | 0.211665425 |
| Shenyang | 0.193133263 | 0.161870655 |
| Dalian | 0.192466245 | 0.161401839 |
| Changchun | 0.21684816 | 0.178204781 |
| Harbin | 0.153156437 | 0.13281497 |
| Shanghai | 0.090138664 | 0.082685503 |
| Nanjing | 0.387868123 | 0.279470446 |
| Hangzhou | 0.137182809 | 0.120633911 |
| Ningbo | 0.235714301 | 0.190751455 |
| Hefei | 0.152769096 | 0.132523588 |
| Fuzhou | 0.107486074 | 0.097054109 |
| Xiamen | 0.167147792 | 0.143210477 |
| Nanchang | 0.278882861 | 0.218067557 |
| Jinan | 0.154347436 | 0.133709688 |
| Qingdao | 0.192033205 | 0.161097194 |
| Zhengzhou | 0.24947193 | 0.199661892 |
| Wuhan | 0.524183519 | 0.343911027 |
| Changsha | 0.27962364 | 0.21852022 |
| Guangzhou | 0.15515039 | 0.134311854 |
| Shenzhen | 0.446092041 | 0.308481084 |
| Nanning | 0.154882855 | 0.134111312 |
| Hainan | 0.267217782 | 0.210869659 |
| Chongqing | 0.240975096 | 0.194182056 |
| Chengdu | 0.291325083 | 0.22560166 |
| Guiyang | 0.399272291 | 0.285342812 |
| Kunming | 1.133531224 | 0.531293478 |
| Lhasa | 2.389924935 | 0.705008217 |
| Xi'an | 0.070399313 | 0.065769206 |
| Lanzhou | 0.323647184 | 0.244511671 |
| Xining | 0.257606402 | 0.204838654 |
| Yinchuan | 0.201130455 | 0.167450966 |
| Urumqi | 0.221941302 | 0.181630085 |
| Average | 0.316685494 (0.231682989) | 0.207391972 |

Figure 1 shows that from 2012-2018, the ratios of range to the maximum blood donation quantities are predominantly clustered around 0.1, while the ratios of range to the minimum donation quantities are mainly concentrated around 0.25.

**Figure 1.**
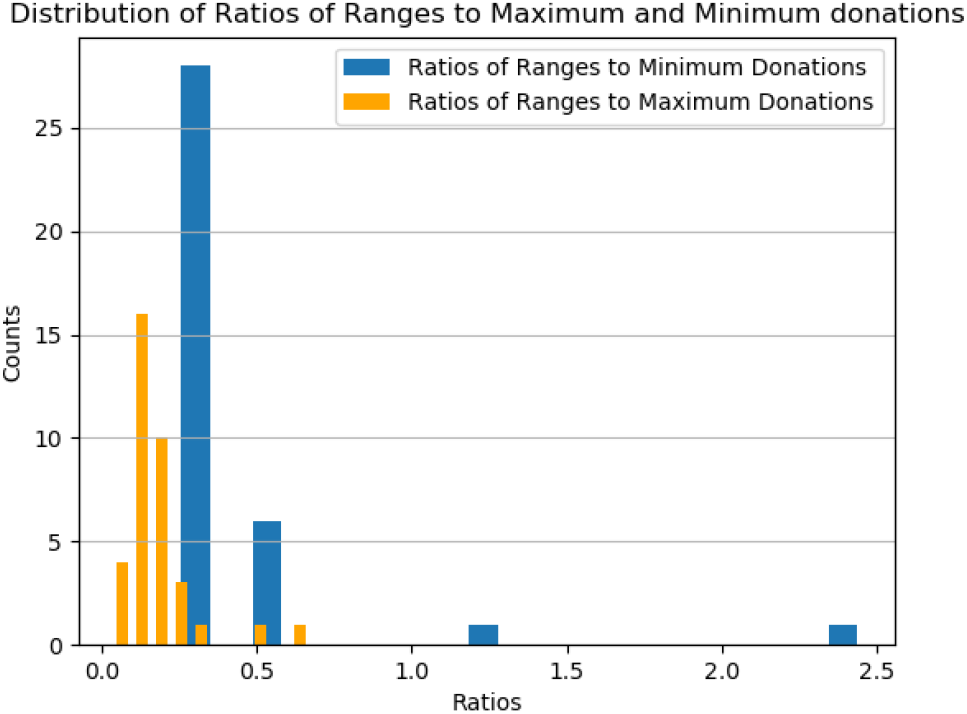
In this figure, the ratios of range to maximum donation quantity for the 36 cities are mostly clustered around 0.1, while the ratios of range to minimum donation quantity are mainly concentrated around 0.25.

### 2.6 Combination of the Methods above

Our method is illustrated in Figure 2, and it involves two approaches for predicting blood donation quantities: the grey prediction model and the linear regression models. The grey prediction model utilizes historical blood donation data from 2012 to 2018 exclusively, and it also makes forecasts for resident population and GDP for the period of 2023-2026, based on data from 2012 to 2021. On the other hand, the linear models use the predicted resident population and GDP as variables to forecast blood donation quantities for the same period. To illustrate the predicted blood donation trends from 2023 to 2026, we present these forecasts using box plots generated by the linear models. For the purposes of this study, we will use the term “variables” to refer to GDP, blood donation quantity, and resident population in each city.

**Figure 2.**
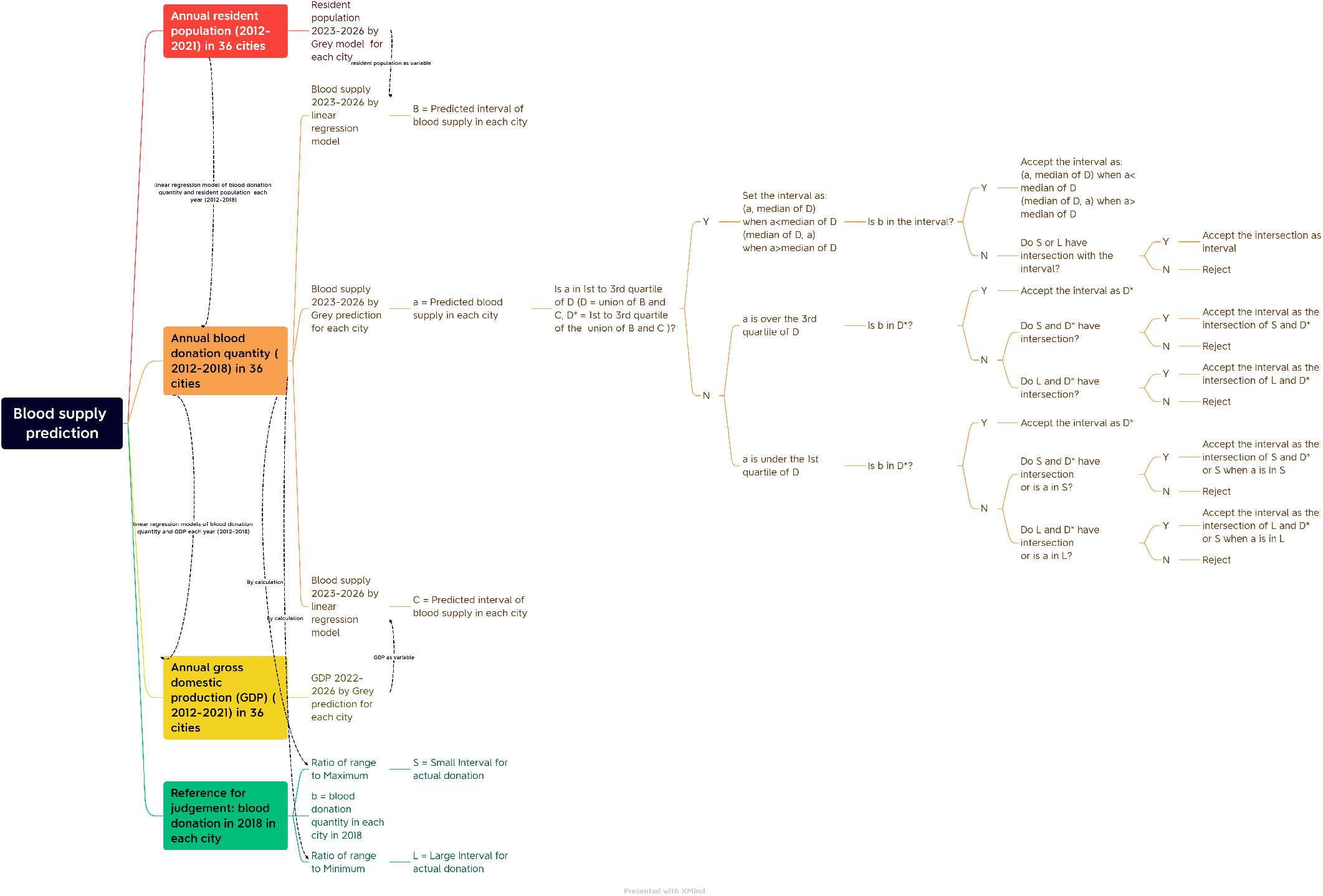
General description of the method

Blood donation quantities for the period 2019-2026 are calculated using prediction models, either the fractional grey prediction model or the linear regression model. However, for the updated information, we preliminarily rely on the results generated by the linear regression models (*D* in Figure 2). This is because the actual data of resident population and GDP had been updated to 2021, while the actual data of blood donation had been updated to 2018. Nonetheless, these predictions have limitations as they can have high errors, resulting in relatively wide predicted intervals. Thus, to enhance precision, we prefer narrower intervals in our predictions. Furthermore, the blood donation quantities predicted by the grey prediction model (element *a* in Figure 2) do not always fall between the interval given by the 1st quartile and 3rd quartile given by the linear regression models (*D*\* in Figure 2). It makes us difficult to judge which preliminary prediction is more close to the most probable blood donation quantity.

To make a final decision on the preliminary results, we need to compare the preliminary predictions with reference assessment interval generated by the latest historical records. However, as of now (in 2023), the latest historical records are from 2018 (*b* in Figure 2), which is far from our prediction period of 2023-2026. Hence, it is necessary to establish principles for selecting upper and lower bounds for the forecasted blood donation quantity. These principles are given below:

1. Cautiousness. We conservatively estimate blood donation quantity to be close to the first quartile of *D*, since blood is a scarce resource.
2. New information priority. We primarily rely on the results generated by the linear regression models when the blood donation quantity predicted by the fractional grey model (*a*) falls outside the interval given by the 1st quartile and 3rd quartile given by the linear regression models (*D*\*).
3. Consistency. This item needs more words to describe.
  (a) Preliminary decisions: If *a* falls within *D*\*, we preliminarily decide that the upper and lower bounds for the forecasted blood donation quantity are given by the 2nd quartile of *D* and *a*. If *a* is larger than the 3rd quartile of *D*, we preliminarily decide that the endpoints of the interval for the forecasted blood donation quantity are the 1st quartile and 2nd quartile of *D* If *a* is smaller than the 1st quartile of *D*, we preliminarily decide that the endpoints of the interval for the forecasted blood donation quantity are the 1st quartile and 2nd quartile of *D* and take the union with *a*.
  (b) Final decisions: To evaluate whether the preliminary predictions are referable and make a final decision of the predicted intervals, we introduce historical records in 2018 (*b*). We construct reference assessment interval around *b* with the error boundary *±*0.1 (reference assessment interval *S*) and *±*0.25 (reference assessment interval *L*).
    i. If *b* falls within the preliminary predicted interval, we accept the preliminarily predicted interval to be the predicted interval.
    ii. If *b* falls out of the preliminary predicted interval, we calculate the intersection of *L* (or *S*) and the sets which we preliminarily predicted:
      A. If the intersection is not null set, and the calculation result is not *a*, we accept the intersection as the predicted interval.
      B. If intersection is not null set, and the calculation result is *a*, we accept *L* (or *S*) to be the predicted interval.
      C. If the intersection is null set, we reject the preliminary prediction to be the predicted interval.

We utilize these principles to establish upper and lower bounds for our predicted intervals. The reason we often select the 2nd quartile as the bounds in our preliminary decision is that it typically occurs more frequently. When reading the principles, readers can consult Figure 2 in the meantime. It’s worth noting that different error margins produces different referable results.

We also record each of the *r*\* in different fractional grey prediction models. The fractional order of the fractional grey model indicates the anticipated trend of the city. A high fractional order indicates that the system exhibits long-term memory, which means that past events have a significant influence on future behavior.

## 3 Results

Following the principles of judgment outlined in section 2, we figured out the predicted intervals of the blood donation quantity in each city in 2023-2026. The predictions of the blood donation quantity in 2023-2026 under different error boundaries are shown in Table 6 and Table 7. Additionally, the predicted blood donation quantities generated by the grey model and the linear regression models are illustrated in supplementary materials in figures 9.

**Table 6:** Referable predictions (U) under the error boundary of *±*0.25

| City | 2023 | 2024 | 2025 | 2026 | Number of referable predictions |
| --- | --- | --- | --- | --- | --- |
| Beijing | (538360.81,597141.86) | (535990.87,624054.27) | (533703.85,632596.94) | (531515.09,632596.94) | 4 |
| Tianjin | (319560.62,327024.55) | (318494.84,334162.99) | (317478.49,341102.58) | (316515.06,347844.09) | 4 |
| Shijiazhuang | N | N | N | N | 0 |
| Taiyuan | (149443.07,167017.04) | (155178.41,176778.77) | (161463.25,187946.6) | (168342.07,200922.09) | 4 |
| Huhot | (57524.9,59588.75) | N | N | N | 1 |
| Shenyang | (203032.28,211074.9) | (209161.84,216467.71) | (215794.54,221595.07) | (221811.37,226100.23) | 4 |
| Dalian | (123749.43,153057.01) | (123340.53,153057.01) | (122929.41,153057.01) | (122525.24,153057.01) | 4 |
| Changchun | (195707.2,212882.64) | (197267.69,214277.5) | (198587.6,214277.5) | (199754.06,214277.5) | 4 |
| Harbin | (198591.22,204249.49) | (198591.22,203644.96) | (198591.22,203061.99) | (198591.22,202504.74) | 4 |
| Shanghai | (480522.24,500640.36) | (483933.02,499669.01) | (486992.11,498603.23) | (489726.11,497481.63) | 4 |
| Nanjing | (285680.81,291492.57) | (288335.94,304709.91) | (290491.72,318645.45) | (292224.9,328059.06) | 4 |
| Hangzhou | (236734.26,292056.97) | (236721.63,292056.97) | (236566.03,292056.97) | (236304.56,292056.97) | 4 |
| Ningbo | N | N | N | N | 0 |
| Hefei | (180826.6,208833.66) | (183616.54,208833.66) | (186397.25,208833.66) | (189163.85,208833.66) | 4 |
| Fuzhou | (130805.46,199373.08) | (130244.93,201659.4) | (129728.96,203830.77) | (129251.9,205890.59) | 4 |
| Xiamen | (91097.14,93681.47) | N | N | N | 1 |
| Nanchang | (168982.7,185643.18) | (175409.26,192899.86) | (182400.66,200632.86) | (189997.7,208871.57) | 4 |
| Jinan | (213519.82,233348.64) | (213425.97,245681.59) | (213229.98,256648.38) | (212957.95,256648.38) | 4 |
| Qingdao | (196344.18,232730.0) | (197448.91,232730.0) | (198319.75,232730.0) | (198993.09,232730.0) | 4 |
| Zhengzhou | (297402.59,311823.12) | (311659.66,326831.48) | (326687.85,342853.85) | (341928.94,359993.26) | 4 |
| Wuhan | (305227.29,313169.57) | (305251.06,341515.94) | (305010.71,367589.38) | (304588.31,367589.38) | 4 |
| Changsha | (274326.7,277180.75) | (275676.96,289412.97) | (276672.48,285230.51) | (277377.96,302703.15) | 4 |
| Guangzhou | (486743.81,558165.02) | (512457.87,560790.82) | (539369.41,562902.64) | (564571.56,567543.18) | 4 |
| Shenzhen | (296905.4,328960.0) | (301479.6,328960.0) | (305453.34,328960.0) | (308896.76,328960.0) | 4 |
| Nanning | (167178.99,198004.1) | (171704.23,204584.01) | (176133.8,211323.33) | (180469.76,218231.91) | 4 |
| Hainan | (104377.5,169410.54) | (103312.62,170718.95) | (102252.26,172001.41) | (101193.28,173255.28) | 4 |
| Chongqing | (628277.34,648021.06) | N | N | N | 1 |
| Chengdu | (412816.07,439637.5) | (423399.49,439637.5) | (433494.38,439637.5) | N | 3 |
| Guiyang | (159325.42,161607.0) | (164060.03,168489.91) | (168142.68,176093.56) | (172047.7,180747.5) | 4 |
| Kunming | (213242.75,217163.47) | (228108.54,233278.33) | (245635.73,253184.89) | (262798.16,273294.62) | 4 |
| Lhasa | N | N | N | N | 0 |
| Xi'an | (293874.52,307506.28) | (312129.87,326804.0) | (331180.44,347491.91) | (351060.63,359600.0) | 4 |
| Lanzhou | N | N | N | N | 0 |
| Xining | N | N | N | N | 0 |
| Yinchuan | (74262.71,91576.94) | (73942.55,91576.94) | (73649.09,91576.94) | (73378.35,91576.94) | 4 |
| Urumqi | N | N | N | N | 0 |
| Total | 30 | 27 | 27 | 26 | 110 |

**Table 7:** Referable predictions (U) within the error boundary of *±*0.1

| City | 2023 | 2024 | 2025 | 2026 | Number of referable predictions |
| --- | --- | --- | --- | --- | --- |
| Beijing | (538360.81,556685.31) | (535990.87,556685.31) | (533703.85,556685.31) | (531515.09,556685.31) | 4 |
| Tianjin | N | N | (317478.49,317784.39) | (316515.06,317784.39) | 2 |
| Shijiazhuang | N | N | N | N | 0 |
| Taiyuan | (166956.52,167017.04) | (166956.52,176778.77) | (161463.25,187946.6) | (168342.07,200922.09) | 4 |
| Huhot | N | N | N | N | 0 |
| Shenyang | (203032.28,211074.9) | (209161.84,213372.66) | N | N | 2 |
| Dalian | (123749.43,134690.17) | (123340.53,134690.17) | (122929.41,134690.17) | (122525.24,134690.17) | 4 |
| Changchun | N | N | N | N | 0 |
| Harbin | N | N | N | N | 0 |
| Shanghai | (480522.24,500640.36) | (483933.02,499669.01) | (486992.11,498603.23) | (489726.11,497481.63) | 4 |
| Nanjing | (285680.81,288691.98) | (288335.94,288691.98) | N | N | 2 |
| Hangzhou | (236734.26,257010.14) | (236721.63,257010.14) | (236566.03,257010.14) | (236304.56,257010.14) | 4 |
| Ningbo | N | N | N | N | 0 |
| Hefei | (180826.6,183773.62) | (183616.54,183773.62) | N | N | 2 |
| Fuzhou | (130805.46,199373.08) | (130244.93,201659.4) | (129728.96,203830.77) | (129251.9,205890.59) | 4 |
| Xiamen | N | N | N | N | 0 |
| Nanchang | (168982.7,185643.18) | (175409.26,190476.11) | (182400.66,190476.11) | (189997.7,190476.11) | 4 |
| Jinan | (213519.82,225850.57) | (213425.97,225850.57) | (213229.98,225850.57) | (212957.95,225850.57) | 4 |
| Qingdao | (196344.18,204802.4) | (197448.91,204802.4) | (198319.75,204802.4) | (198993.09,204802.4) | 4 |
| Zhengzhou | N | (323151.3,326831.48) | (326687.85,342853.85) | (341928.94,359993.26) | 3 |
| Wuhan | (305227.29,313169.57) | (305251.06,323478.65) | (305010.71,323478.65) | (304588.31,323478.65) | 4 |
| Changsha | (274326.7,277180.75) | (275676.96,284463.64) | (276672.48,284463.64) | (277377.96,284463.64) | 4 |
| Guangzhou | (486743.81,558165.02) | (512457.87,560790.82) | (539369.41,562902.64) | (564571.56,567543.18) | 4 |
| Shenzhen | N | N | N | N | 0 |
| Nanning | (179850.6,198004.1) | (171704.23,204584.01) | (176133.8,211323.33) | (180469.76,218231.91) | 4 |
| Hainan | (104377.5,169410.54) | (103312.62,170718.95) | (102252.26,172001.41) | (101193.28,173255.28) | 4 |
| Chongqing | N | N | N | N | 0 |
| Chengdu | N | N | N | N | 0 |
| Guiyang | N | N | N | N | 0 |
| Kunming | N | (231654.64,233278.33) | (245635.73,253184.89) | (262798.16,273294.62) | 3 |
| Lhasa | N | N | N | N | 0 |
| Xi'an | (293874.52,307506.28) | (312129.87,316448.0) | N | N | 2 |
| Lanzhou | N | N | N | N | 0 |
| Xining | N | N | N | N | 0 |
| Yinchuan | (74262.71,80587.7) | (73942.55,80587.7) | (73649.09,80587.7) | (73378.35,80587.7) | 4 |
| Urumqi | N | N | N | N | 0 |
| Total | 19 | 21 | 18 | 18 | 76 |

We tallied the number of successful predictions for each city and year. As shown in Table 6, the error boundary is *±*0.25. Our predictions have been proven referable in over two-thirds of the cities. This means that under the error boundary of *±*0.25, over two-thirds of the cities can refer to our prediction. It is hypothesized that the blood donation intervals in 2023-2026 will be wide in each city. This judgement is optimistic.

We also have pessimistic judgement. Regard the blood donation intervals in each city to be narrow in 2023-2026. At this circumstances, the error boundary is *±*0.1, and the predictions have been proven referable in over half of the cities. The results are presented in Table 7.

Consulting Tables 6-7 and the resident population in Annual Statistical Review, it is found that most of the cities have the resident population around 8-15 million, the predicted annual donation quantities are around 200,000 U - 300,000 U. For the cities where the resident populations are around 20 million, the future annual blood donation quantity will be around 400,000 U. If a city where the resident population is over 30 million the future annual blood donation quantity will be around 600,0000 U. It can be concluded that when our prediction method works in that city, the predicted average annual blood donation quantity per person can be estimated as around 20 U per thousand persons. The donation quantity in these cities can be estimated according to the resident population. The actual resident population in 2018 and the blood donation quantity are shown in section 9, the supplementary materials Table 8, and the predicted resident population in 2023 to 2026 are also shown in Table 9.

**Table 8:** The actual resident population and blood donation quantity in 2018

| City | Resident Population (10,000 persons) | Blood Donation Quantity (U) | Donation Quantity per 1000 persons (U) |
| --- | --- | --- | --- |
| Beijing | 2154.2 | 506077.55 | 23.49 |
| Tianjin | 1559.6 | 288894.9 | 18.52 |
| Shijiazhuang | 1095.16 | 321824 | 29.39 |
| Taiyuan | 442.15 | 185507.25 | 41.96 |
| Huhot | 312.6 | 47671 | 15.25 |
| Shenyang | 831.6 | 193975.15 | 23.33 |
| Dalian | 700.2 | 122445.61 | 17.49 |
| Changchun | 767.4 | 171422 | 22.34 |
| Harbin | 1085.8 | 264788.3 | 24.39 |
| Shanghai | 2423.78 | 456390 | 18.83 |
| Nanjing | 843.62 | 262447.25 | 31.11 |
| Hangzhou | 980.6 | 233645.58 | 23.83 |
| Ningbo | 820.2 | 128608.77 | 15.68 |
| Hefei | 808.7 | 167066.93 | 20.66 |
| Fuzhou | 774 | 137484.46 | 17.76 |
| Xiamen | 411 | 74945.18 | 18.23 |
| Nanchang | 554.55 | 173160.1 | 31.23 |
| Jinan | 746.04 | 205318.7 | 27.52 |
| Qingdao | 939.48 | 186184 | 19.82 |
| Zhengzhou | 1013.6 | 359057 | 35.42 |
| Wuhan | 1108.1 | 294071.5 | 26.54 |
| Changsha | 815.47 | 258603.31 | 31.71 |
| Guangzhou | 1490.44 | 531122.63 | 35.64 |
| Shenzhen | 1343.88 | 263168 | 19.58 |
| Nanning | 725.41 | 199834 | 27.55 |
| Hainan | 934.32 | 162029 | 17.34 |
| Chongqing | 3101.79 | 518416.85 | 16.71 |
| Chengdu | 1633 | 351710 | 21.54 |
| Guiyang | 488.19 | 144598 | 29.62 |
| Kunming | 685 | 257394.05 | 37.58 |
| Lhasa | 55.89 | 1705.2 | 3.05 |
| Xi'an | 1000.37 | 287680 | 28.76 |
| Lanzhou | 375.36 | 80696.5 | 21.5 |
| Xining | 237.11 | 65851 | 27.77 |
| Yinchuan | 225.06 | 73261.55 | 32.55 |
| Urumqi | 350.58 | 73603 | 20.99 |

**Table 9:** The predicted resident population in 2023-2026

| City | 2023 | 2024 | 2025 | 2026 |
| --- | --- | --- | --- | --- |
| Beijing | 2159.15 | 2151.58 | 2144.17 | 2137 |
| Tianjin | 1450.62 | 1445.01 | 1439.67 | 1434.6 |
| Shijiazhuang | 1119.04 | 1118.83 | 1118.07 | 1116.89 |
| Taiyuan | 595.41 | 626.79 | 661.17 | 698.8 |
| Huhhot | 368.77 | 378.91 | 389.3 | 399.93 |
| Shenyang | 953.61 | 976.26 | 999.81 | 1024.26 |
| Dalian | 757.24 | 762.06 | 766.22 | 769.8 |
| Changchun | 979.73 | 1018.22 | 1059.68 | 1104.26 |
| Harbin | 1032.12 | 1027.82 | 1023.82 | 1020.11 |
| Shanghai | 2465.11 | 2459.97 | 2454.33 | 2448.4 |
| Nanjing | 1006.62 | 1044.58 | 1085.8 | 1130.47 |
| Hangzhou | 1387.67 | 1479.81 | 1581.61 | 1693.99 |
| Ningbo | 1031.35 | 1072.41 | 1115.73 | 1161.37 |
| Hefei | 1043.65 | 1104.91 | 1173.74 | 1250.97 |
| Fuzhou | 868.55 | 881.06 | 892.94 | 904.2 |
| Xiamen | 598.35 | 636.75 | 678.57 | 724.08 |
| Nanchang | 702.3 | 737.46 | 775.7 | 817.26 |
| Jinan | 1051.05 | 1116.3 | 1188.26 | 1267.56 |
| Qingdao | 1057.33 | 1073.04 | 1087.92 | 1102 |
| Zhengzhou | 1421.2 | 1500.24 | 1584.86 | 1675.38 |
| Wuhan | 1473.33 | 1623.29 | 1807.21 | 2032.41 |
| Changsha | 1164.03 | 1241.09 | 1325.52 | 1417.96 |
| Guangzhou | 2123.04 | 2250.79 | 2388.09 | 2535.57 |
| Shenzhen | 2056.24 | 2206.71 | 2368.59 | 2542.66 |
| Nanning | 965.22 | 1009.08 | 1055.4 | 1104.28 |
| Hainan | 1059.49 | 1078.9 | 1098 | 1116.77 |
| Chongqing | 3232.7 | 3240.75 | 3246.5 | 3250.31 |
| Chengdu | 2417.01 | 2576.01 | 2748.3 | 2934.89 |
| Guiyang | 681.2 | 719.79 | 761.54 | 806.65 |
| Kunming | 972.59 | 1063.22 | 1172.23 | 1303.16 |
| Lhasa | 98.19 | 104.24 | 110.66 | 117.47 |
| Xi'an | 1502.74 | 1615.19 | 1738.78 | 1874.53 |
| Lanzhou | 481.71 | 511.05 | 544.76 | 583.45 |
| Xining | 252.05 | 254.49 | 256.77 | 258.88 |
| Yinchuan | 322.36 | 340.73 | 360.47 | 381.67 |
| Urumqi | 434.17 | 449.1 | 464.73 | 481.07 |

We also put the number of referable predictions on a map, see Figure 3. According to Figure 3, the areas with strong reference value in the prediction results are mainly concentrated in the central and eastern parts of China. These areas are economically prosperous and densely populated. This guarantees the blood supply to be stable.

**Figure 3.**
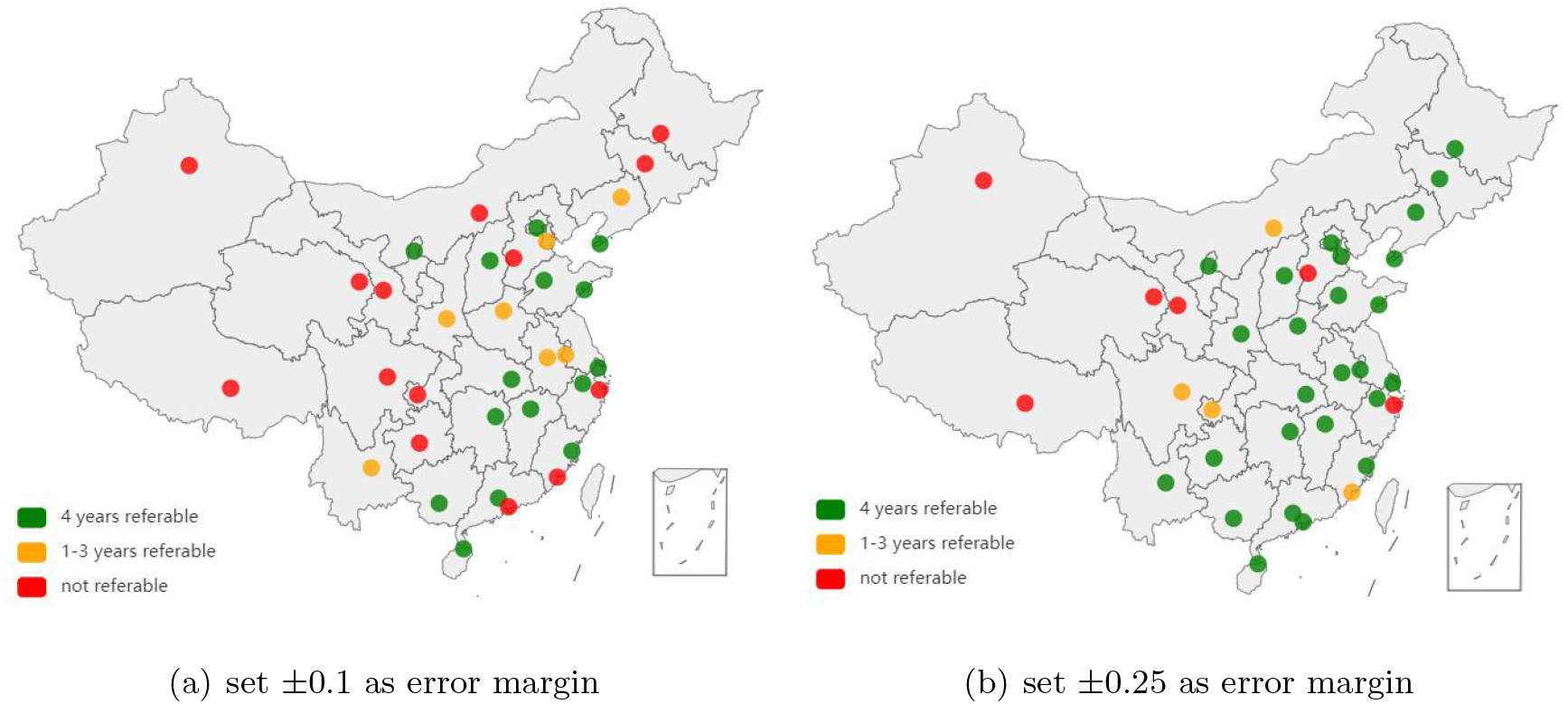
The predictions are acceptable in over half of the cities

So far, we have presented short-term predictions of blood donation quantity in each city. The long-term prediction depends on the fractional order of the fractional grey prediction model. We have already mentioned in section 2 that the fractional order of the fractional grey model indicates the anticipated trend of the city. A high fractional order indicates that the system exhibits long-term memory, which means that past events have a significant influence on future behavior. Figures 4 depict the fractional differential orders for each variable. These figures reveal that the changes in resident population, GDP, and blood donation quantities across cities do not always occur in unison. Figure a and b in Figures 4 suggest that the changes in resident population depend on near-historical data, and the changes in GDP in northeast China depend on early historical data. The fractional orders for each of the grey model are listed in Table 10 in the supplementary materials section 9.

**Table 10:** The fractional order of the fractional grey model in each of the areas

| City | Order for blood donation | Order for resident population | Order for GDP |
| --- | --- | --- | --- |
| Beijing | 0.06 | 0.05 | 0.05 |
| Tianjin | 0.05 | 0.05 | 0.06 |
| Shijiazhuang | 0.05 | 0.05 | 0.05 |
| Taiyuan | 0.06 | 0.05 | 0.05 |
| Huhhot | 0.06 | 0.05 | 0.06 |
| Shenyang | 0.05 | 0.05 | 0.06 |
| Dalian | 0.05 | 0.05 | 0.06 |
| Changchun | 0.06 | 0.05 | 0.05 |
| Harbin | 0.05 | 0.05 | 0.06 |
| Shanghai | 0.05 | 0.05 | 0.05 |
| Nanjing | 0.05 | 0.05 | 0.05 |
| Hangzhou | 0.05 | 0.05 | 0.05 |
| Ningbo | 0.05 | 0.05 | 0.05 |
| Hefei | 0.05 | 0.05 | 0.05 |
| Fuzhou | 0.05 | 0.05 | 0.05 |
| Xiamen | 0.05 | 0.05 | 0.05 |
| Nanchang | 0.06 | 0.05 | 0.05 |
| Jinan | 0.05 | 0.05 | 0.05 |
| Qingdao | 0.05 | 0.05 | 0.05 |
| Zhengzhou | 0.06 | 0.05 | 0.05 |
| Wuhan | 0.05 | 0.06 | 0.05 |
| Changsha | 0.05 | 0.05 | 0.05 |
| Guangzhou | 0.05 | 0.05 | 0.05 |
| Shenzhen | 0.05 | 0.06 | 0.05 |
| Nanning | 0.05 | 0.05 | 0.05 |
| Hainan | 0.06 | 0.05 | 0.06 |
| Chongqing | 0.05 | 0.05 | 0.05 |
| Chengdu | 0.05 | 0.05 | 0.05 |
| Guiyang | 0.05 | 0.05 | 0.05 |
| Kunming | 0.06 | 0.05 | 0.05 |
| Lhasa | 0.06 | 0.06 | 0.05 |
| Xi'an | 0.05 | 0.06 | 0.05 |
| Lanzhou | 0.06 | 0.05 | 0.05 |
| Xining | 0.06 | 0.05 | 0.05 |
| Yinchuan | 0.05 | 0.05 | 0.05 |
| Urumqi | 0.06 | 0.05 | 0.05 |

**Figure 4.**
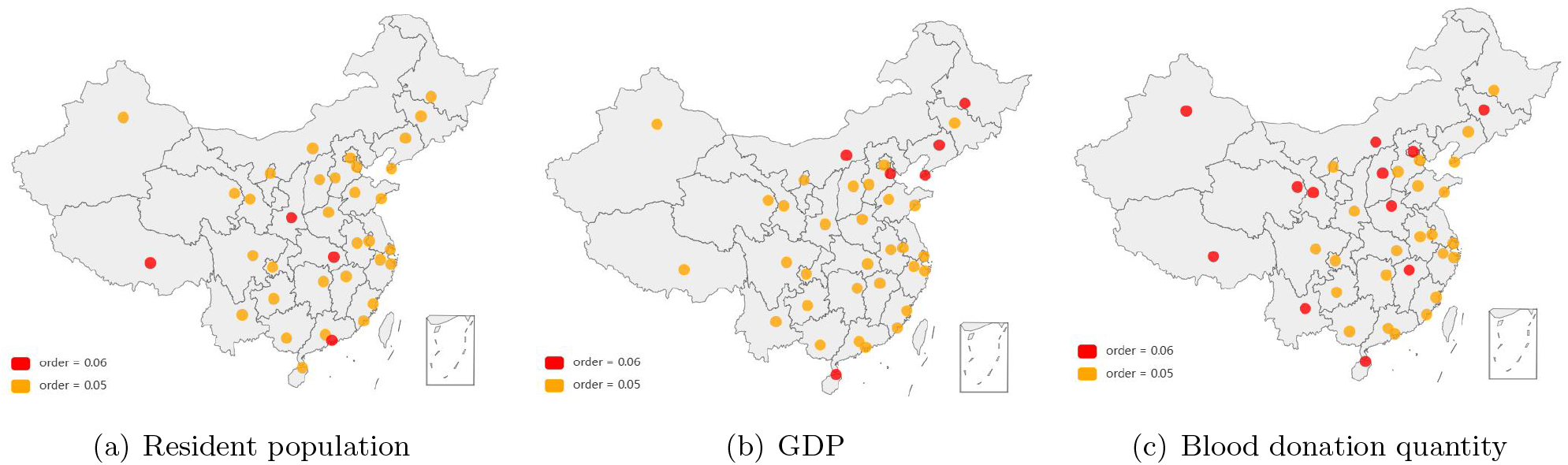
Fractional differential orders of each kinds of variables in each of the areas

## 4 Conclusion and Discussion

Given the scarcity of blood as a resource, our estimation of blood donation quantity must be conservative and risk-averse, as stated in section 2. This approach can help hospitals evaluate their blood use in the near future. For clinical blood use, forecasting the blood donation quantity for the next 1-2 years is sufficient, and given the limitations of the grey prediction model for long-term predictions, it is advisable to adjust the fractional order at the end of each new year. Additionally, historical records that are too far from the timescale of interest should be omitted from the forecasting process. When it comes to policy analysis, forecasting the blood donation quantity for the next 3-5 years is crucial, and obtaining continuous historical records is the best way to achieve accurate forecasting. Unfortunately, due to the pandemic, some historical records were not referable, necessitating the use of two independent approaches to predict future blood donation quantities.

Based on our methods, we can conclude that for the cities where our methods work the blood centers can predict blood supply for the next 1-2 years. After communication with hospitals, blood centers can estimate the number of donors for recruiting, when there is a gap between the forecast supply and clinical needs. We also need to pay attention to the western part of China, where the predictions of blood donation are not quite referable. We find that the economy is underdeveloped and the population density is low in these areas. Comparing the western part of China with the central and eastern parts of China, it is obvious that the central and eastern parts of China are economically prosperous and densely populated. This suggests that optimizing the economic and demographic structure of western China can help accurately predict blood supply and thus optimize the utilization of local blood resources.

The fractional order parameter provides valuable insights into the specific characteristics of each area. It is determined by seeking the best match between the predicted and historical data. When different areas have the same fractional order for a given variable, it suggests that they may share similar developmental patterns. This is helpful for long-term estimation of blood supply. Based on the results found in this study, we should pay much attention to the northeast China, where there will be population loss. This will affect the blood supply in a long run. So far we can use our prediction method to forecast the blood donation intervals in this area, but this method might lose efficacy 5 years later.

## Data Availability

All data produced in the present study are available upon reasonable request to the authors

http://www.stats.gov.cn/sj/ndsj/

## 5 Acknowledgements

This study was supported by Fundamental Research Funds for the Central Universities (3332019170)

## 7 Author Statements

Author contributions: Feng Lin designed the study. Yi Huang, Xu He, Huifang Zhang and Feng Lin collected the data. Zhong Liu supplied the data. Feng Lin, Xu He and Yi Huang analyzed the data. Feng Lin, Xu He and Yi Huang wrote the manuscript. All authors reviewed the manuscript

## 8 Data Statements

All data produced in the present study are available upon reasonable request to the authors

## 9 Supplementary Materials

The actual blood donation quantity and the resident population in 2018 are shown in Table 8

The predicted resident population in 2023-2026 are shown in Table 9

The fractional order of the variables in each of the areas when fractional grey prediction model is used are shown in Table 10.

The blood donation quantity in each of the cities predicted by grey prediction model and the linear model are shown in Figures 9

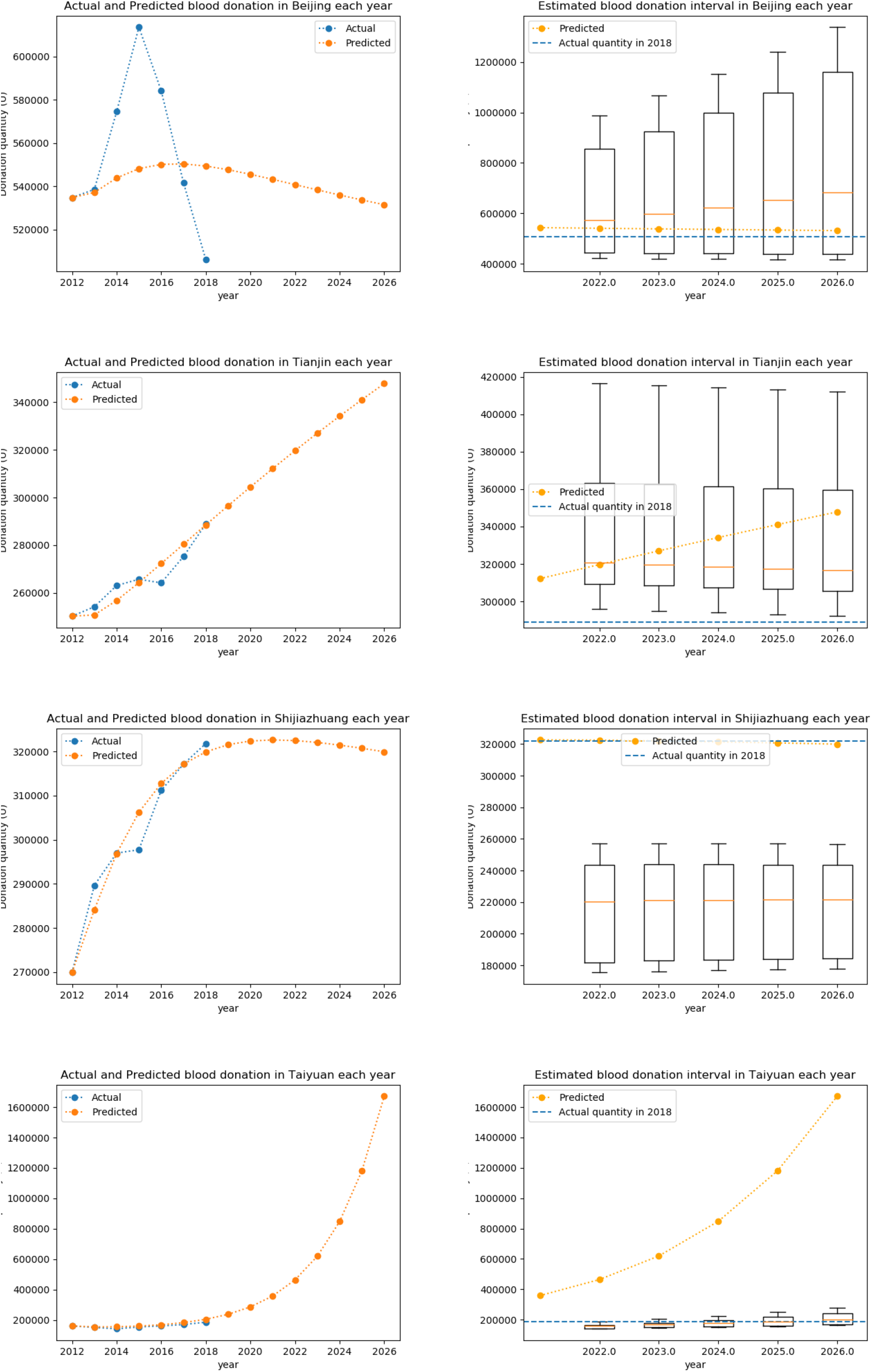

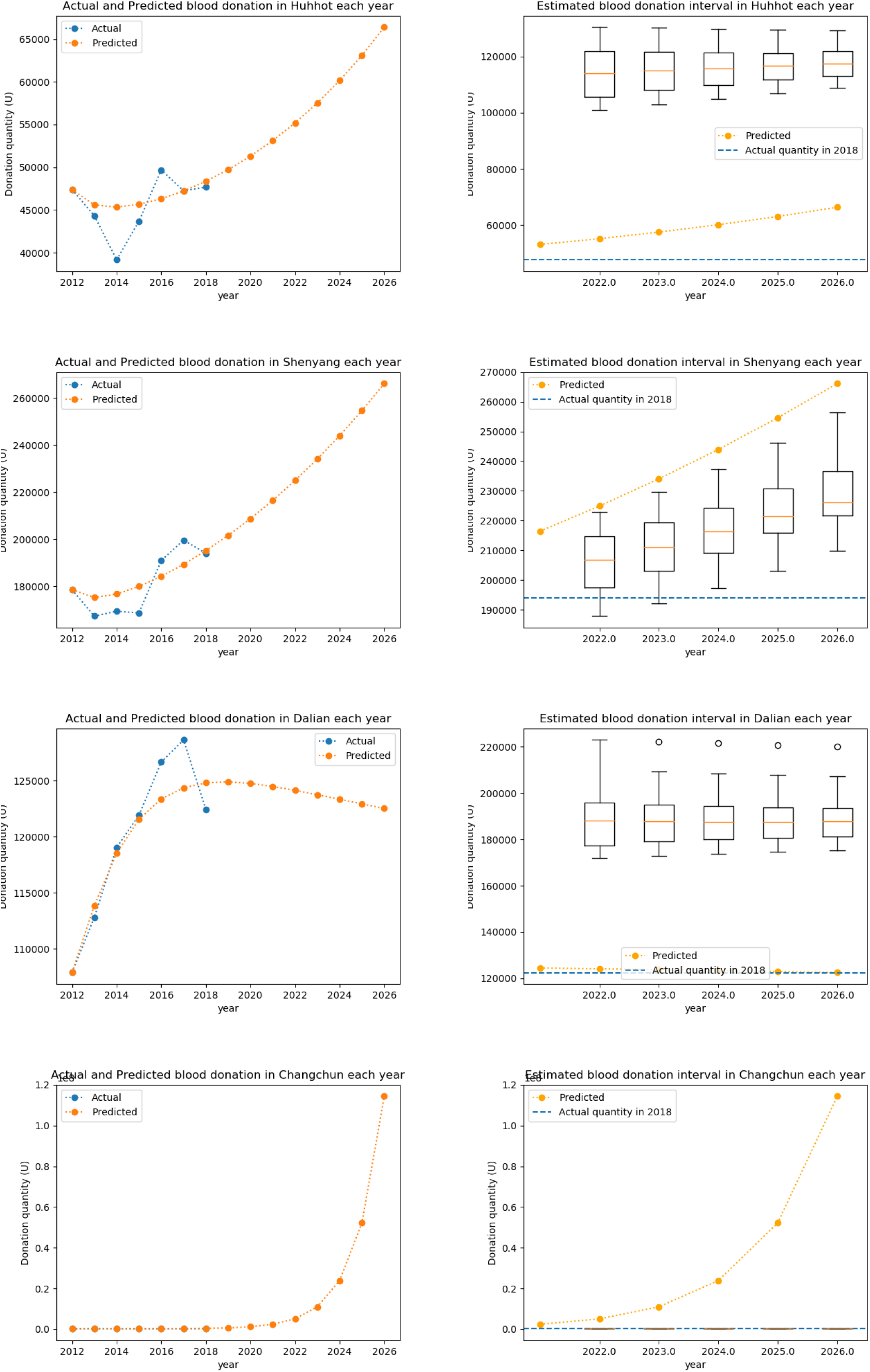

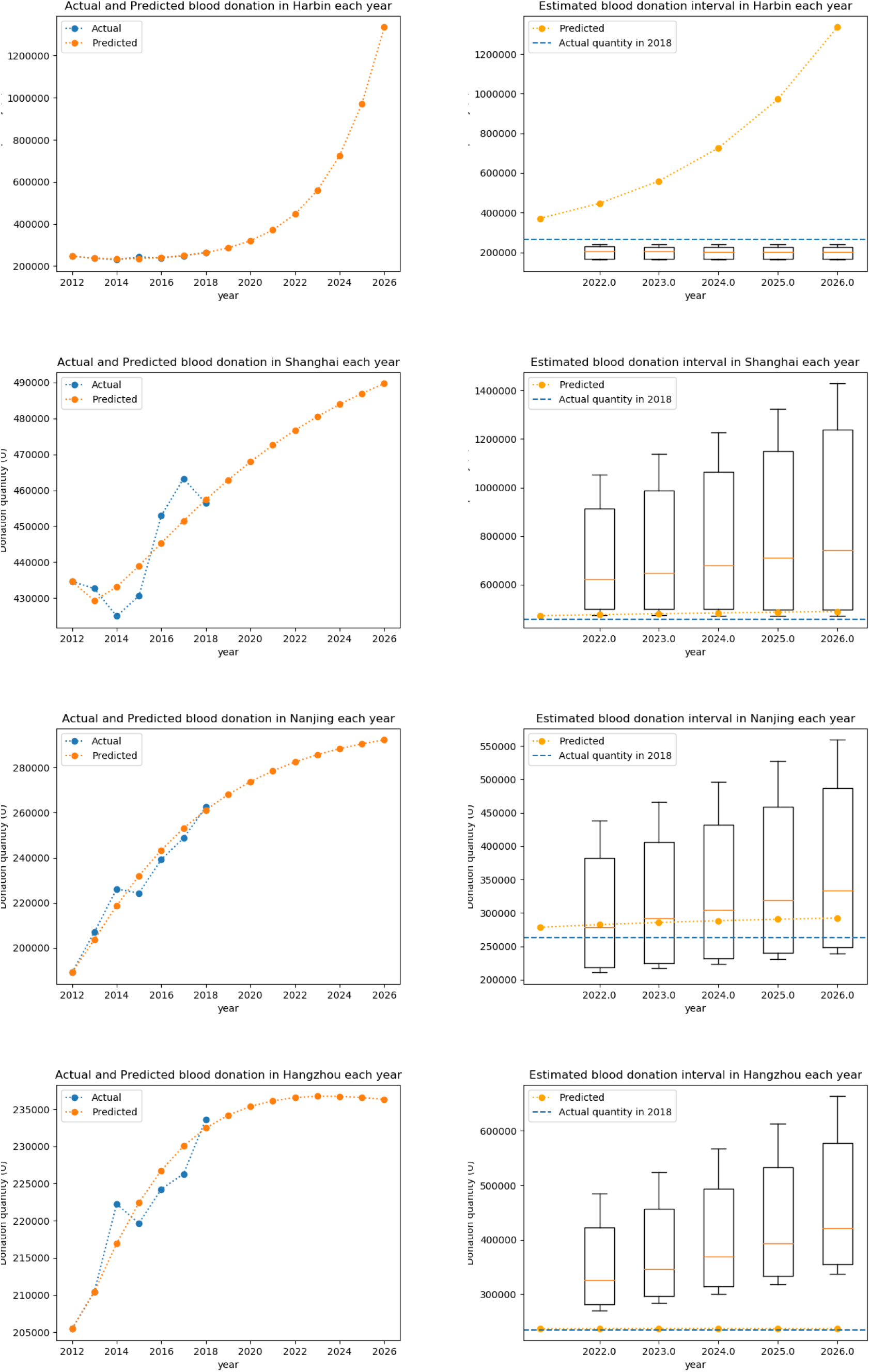

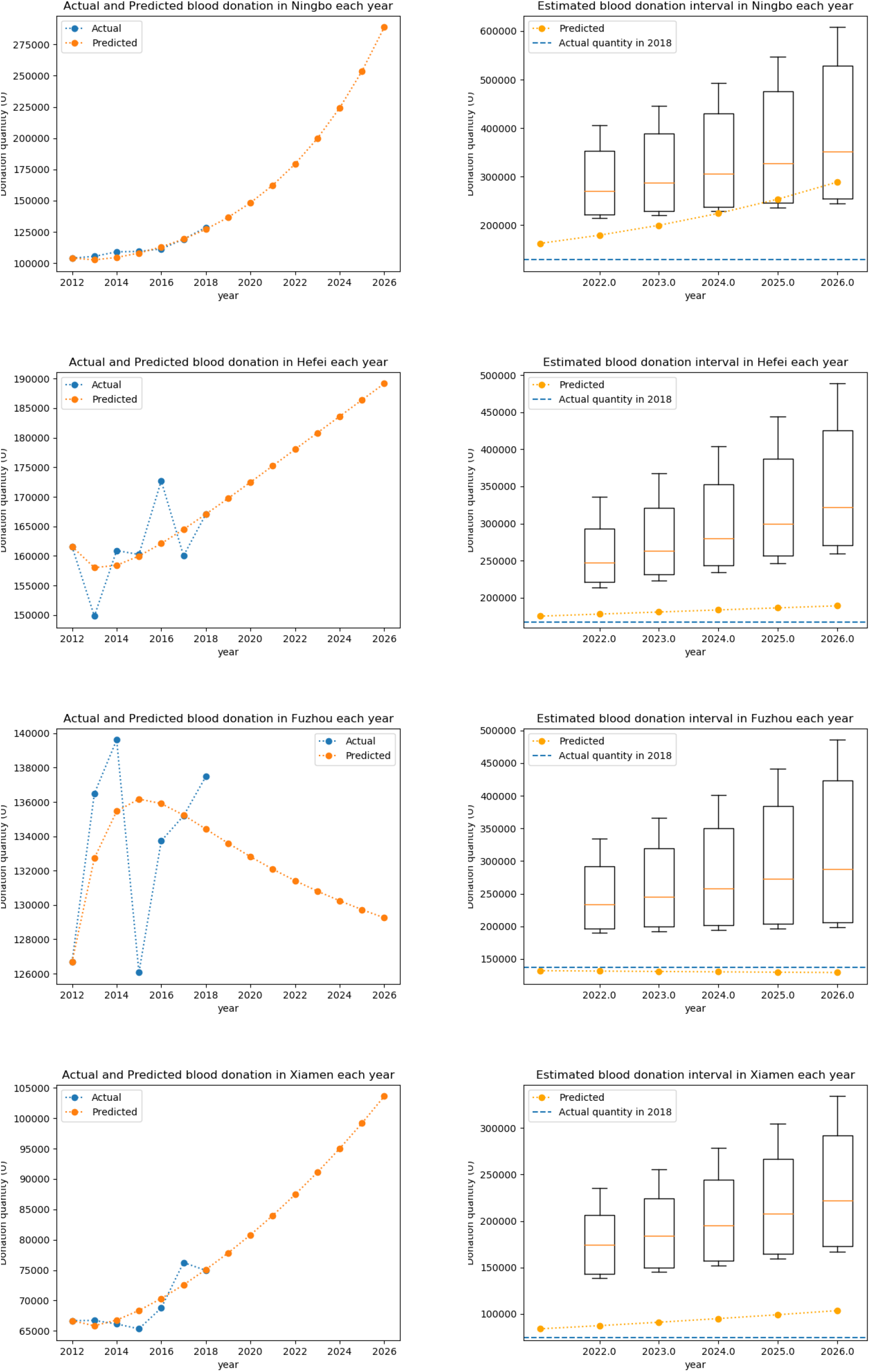

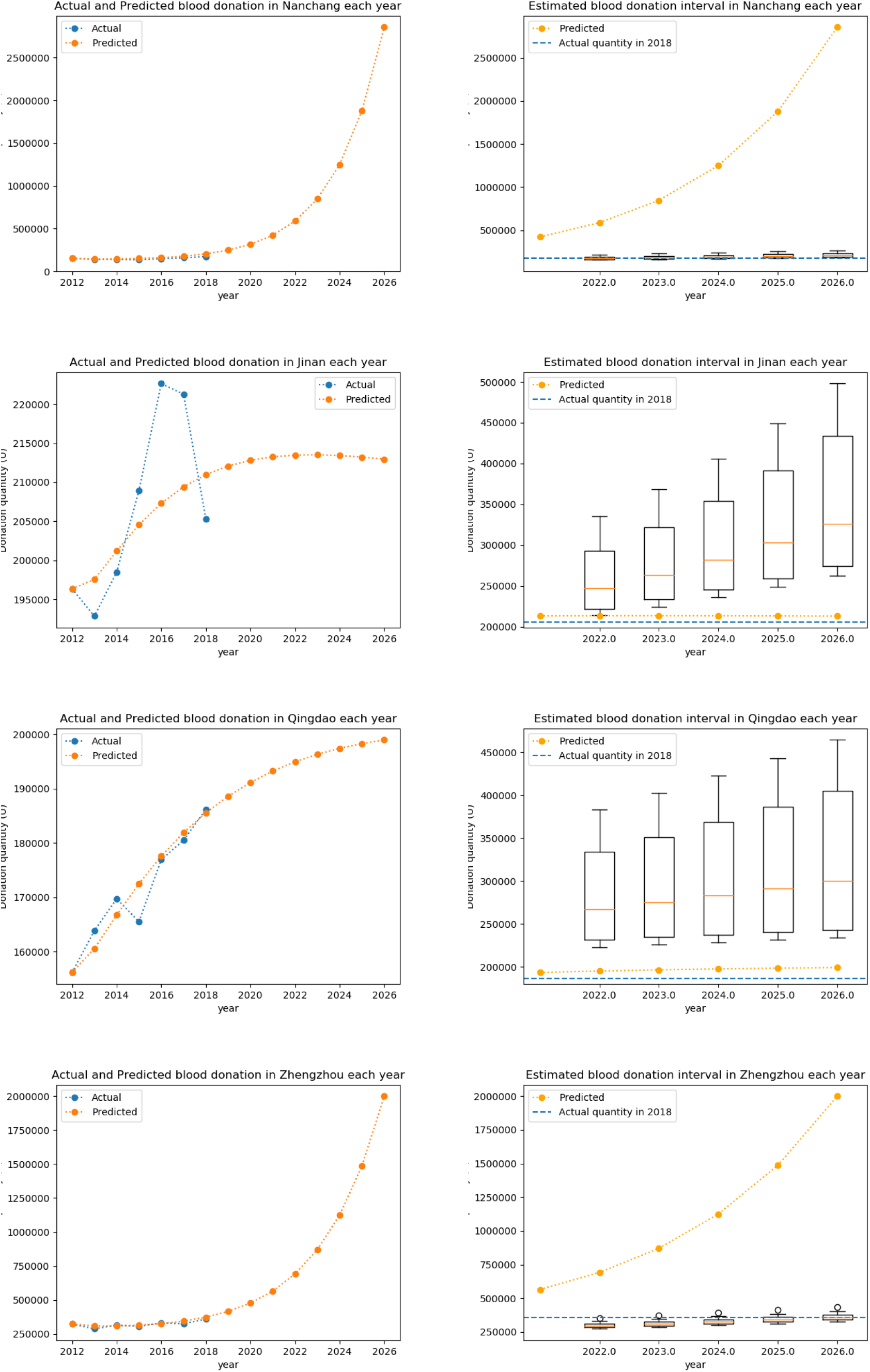

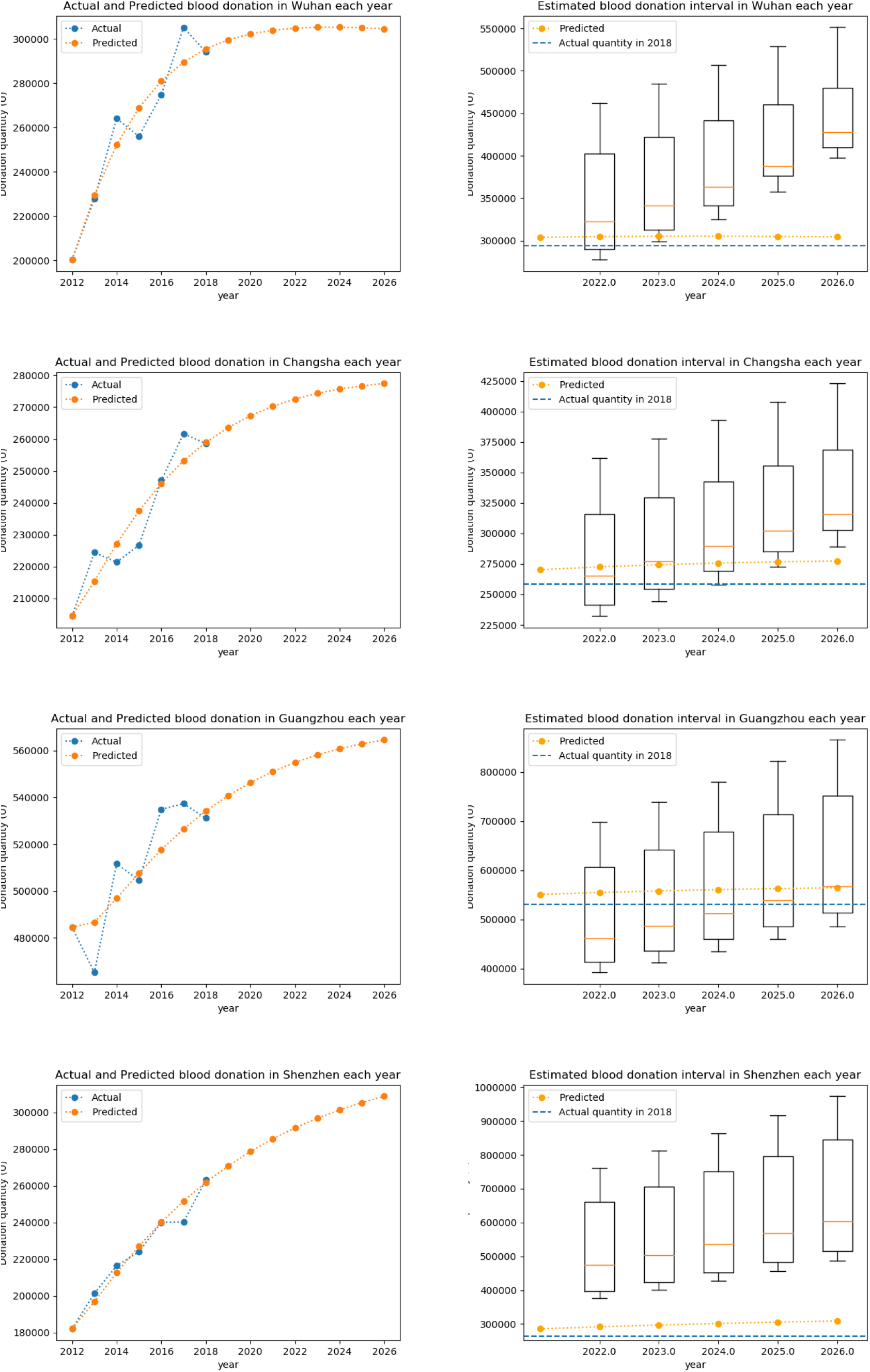

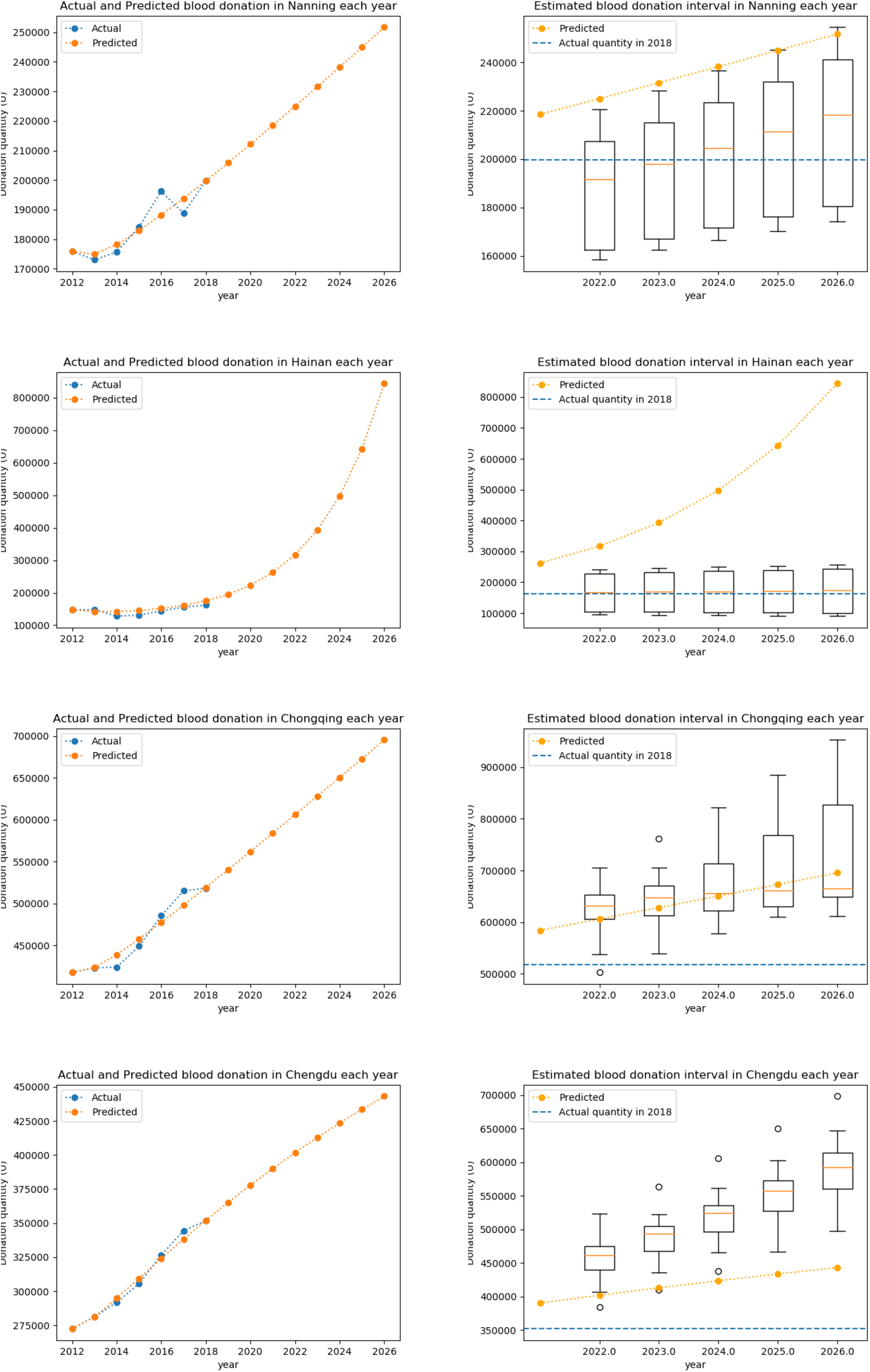

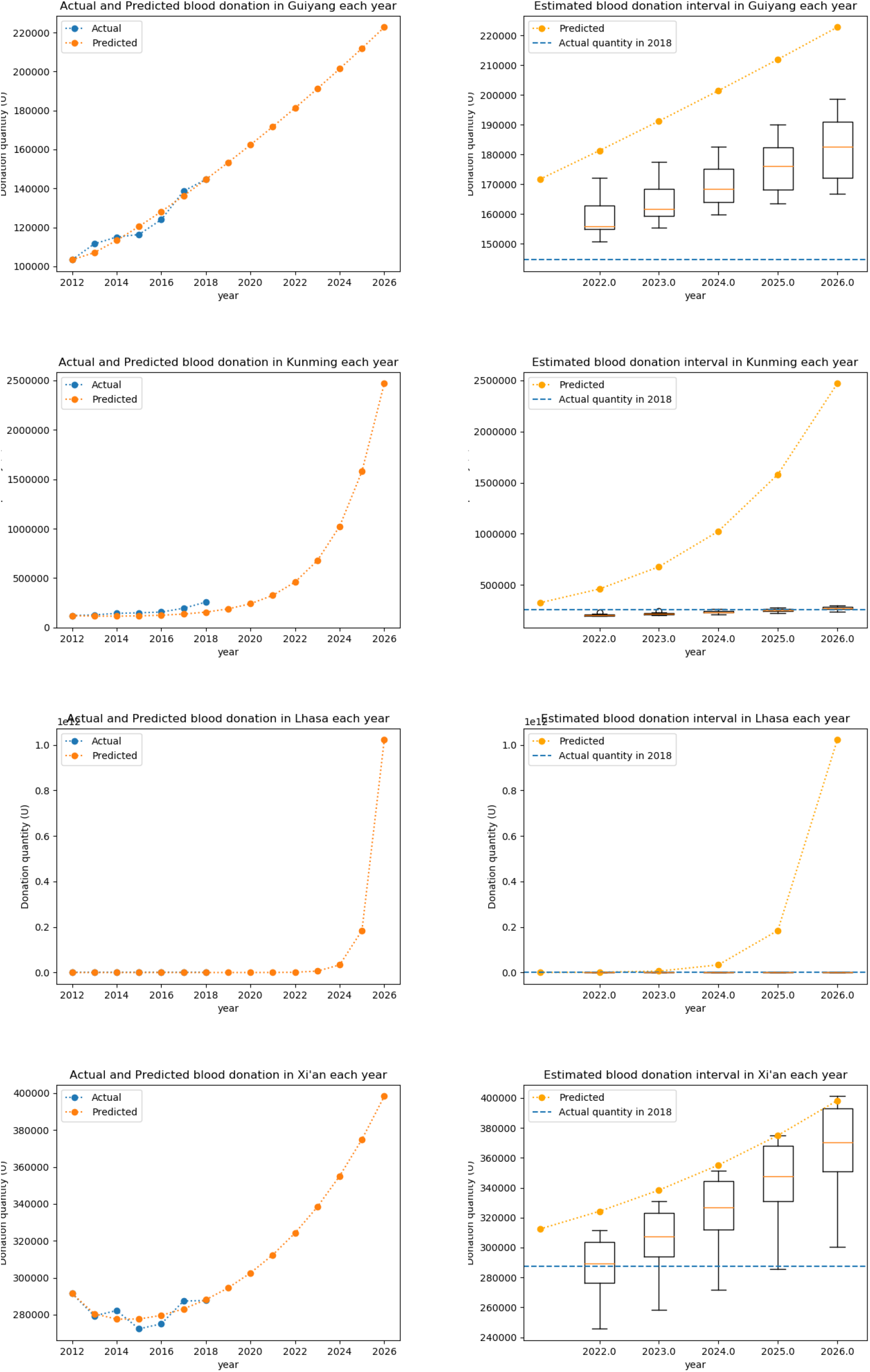

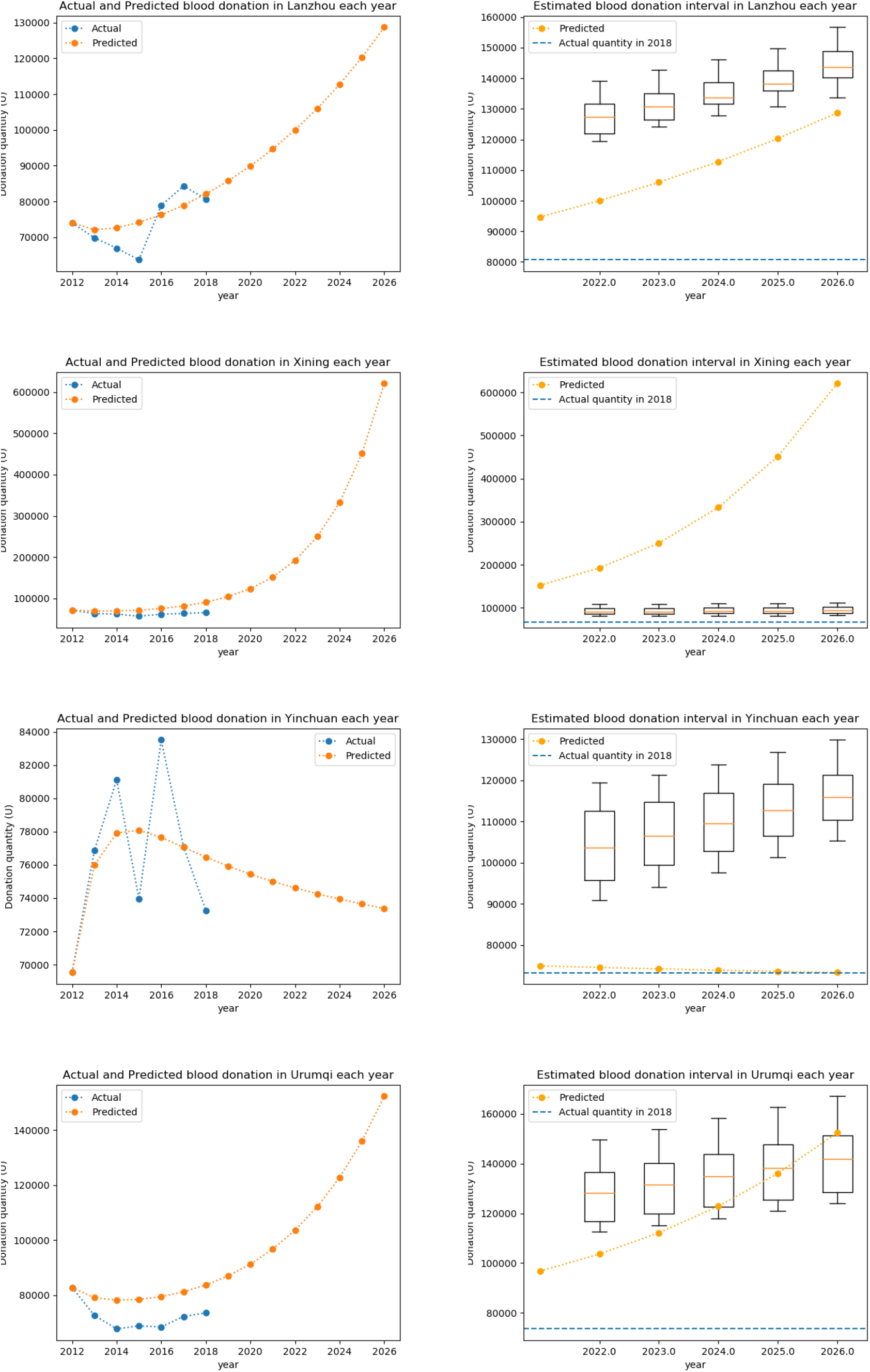

